# Time to early ambulation and its predictors among admitted patients undergoing abdominal surgery in East Gojjam Zone Public Hospitals, Northwest Ethiopia

**DOI:** 10.1101/2025.05.18.25327872

**Authors:** Afework Edmealem, Tiliksew Liknaw, Anteneh Kehaliw Temesgen, Tirusew Wonda, Setarg Ayenew, Bayachew Asmare, Temesgen Ayenew, Bewuketu Zelalem Zeleke, Ayenew Sisay Gebeyew, Addisu Getie

## Abstract

**Introduction:** Early ambulation has demonstrated numerous benefits in the postoperative period. Despite the recognized advantages of early ambulation, there exists considerable variability in the time taken by patients to initiate ambulatory activities. Thus, this study aimed to assess the time to early ambulation and its predictors among patients undergoing abdominal surgery in East Gojjam Zone Public Hospitals, Northwest Ethiopia.

**Methods:** Institutional based prospective follow-up study was conducted among 444 patients undergoing abdominal surgery by using systematic sampling. Patients were followed for 24 hours along chart review and interview. After checking the cox proportional hazard assumptions and model fitness test, cox proportional regression analysis model was conducted by using STATA 17 software. Variables with a P value less than 0.2 in the bivariable analysis were selected as candidates for the multivariable cox proportional regression. An adjusted hazard ratio with its 95% CI was used to show the strength of association and P-value less than 0.05 was used to declare statistical significance.

**Result:** From the total of 444 patients undergoing abdominal surgery and followed for 24 hours, 80.6% (95% CI: 0.77-0.84) of them were ambulated. The incidence rate of early ambulation was 5.64% per 100 patient-hours follow up. The median time to early ambulation was 13 hours. Age (AHR = 0.98; 95%CI: (0.97-0.99)), not having catheter (AHR-1.77; 95% CI: (1.37-2.29)), taking tramadol for antipain (AHR-0.26; 95% CI: (0.15-0.45)), postoperative diastolic blood pressure (AHR - 1.01; 95% CI: (1.00-1.02)), and respiratory rate (AHR-0.81; 95% CI: (0.75-0.87)), intraoperative pulse rate(AHR-0.98, 95% CI: (0.97-0.99)), and intraoperative temperature (AHR-1.50, 95% CI: (1.11-2.03)) were found to be predictors of early ambulation.

**Conclusion:** The median time to early ambulation was 13 hours. Thus, health care providers, particularly nurses and surgeons, should actively promote and facilitate early ambulation within 13 hours following abdominal surgery. Increased age, not having catheter, taking tramadol for antipain, postoperative diastolic blood pressure and respiratory rate, intraoperative pulse rate and temperature were predictors of time to early ambulation. As a result, it is better to use antipains other than tramadol in addition to encouraging early ambulation among aged patients.

## Introduction

Major abdominal surgery is major surgery procedures with abdominal organs[1]. Abdominal surgery is a common and sometimes unavoidable operation in most medical specialties, including general surgery, gynecology, urology, and others. It encompasses a wide range of procedures, including appendectomy, cholecystectomy, and bowel resection, and is performed worldwide on a regular basis either for the purpose of diagnosis or therapeutics[2, 3]. Surgical procedures have increased considerably in recent years with rapid technological developments in the field of health[4]. Major abdominal surgery is among surgical procedures that result in a significant loss of functional capacity and complications in spite of technological advances and progress in patient care[5]. In randomized controlled trials (RCTs), the complication rate after major abdominal surgery has been reported between 30% and 60% [6–9]. The main and most difficult complications are bleeding, organ ischemia, surgical site wound infection, anastomotic leakage and cardiovascular problems[1, 9].

Surgical techniques and perioperative care have significantly improved outcome[10]. The postoperative recovery phase following abdominal surgery is a critical period that significantly impacts patient outcomes, including morbidity, mortality, and healthcare costs[11, 12].

In recent years, as the healthcare field strives to optimize patient recovery and improve outcomes, the implementation of Enhanced Recovery After Surgery (ERAS) protocols has gained attention[10, 11, 13]. One of the key components of ERAS is early ambulation, which refers to the initiation of walking and movement soon after abdominal surgery[14, 15]. According to the ERAS protocol, it is recommended that the patient should stay out of bed for two hours on the day of surgery and six hours a day until discharge in the following days[16].

Early ambulation has demonstrated numerous benefits in the postoperative period. Early postoperative mobilization can potentially prevent surgical complications by avoiding prolonged bed rest[17]. It has been associated with a reduction in the incidence of postoperative complications, such as pneumonia[18], deep vein thrombosis, and surgical site infections [17]. Additionally, early ambulation plays a crucial role in improving respiratory function by preventing atelectasis and promoting effective lung expansion[18–20]. It also stimulates gastrointestinal motility, leading to faster recovery of bowel function and a decreased risk of ileus[21]. Moreover, early ambulation has been shown to contribute to shorter hospital stays[19, 22–24], resulting in reduced healthcare costs[18] and improved sleep quality[25], activities of daily living[26] and patient satisfaction, and reduced the anxiety levels[27]. More interestingly, although early mobilization is inhibited by pain, early mobilization helps to ease the postoperative pain after abdominal surgery[27].

Despite guideline recommendations, studies have shown that patients’ adherence to early mobilization was rather low. For example, a study done on factors associated with early mobilization among colorectal cancer patients after surgery and adherence to ERAS elements in major visceral surgery reported that less than 50% of the patients got out of bed on the first postoperative day, and 20% of patients did not walk in the ward until the fifth postoperative day[28, 29].

Due to this delayed ambulation has resulted in prolonged hospital stays, and higher rates of postoperative complications[30–32]. A systematic review and meta-analysis reported that 5.9% patients undergoing abdominal surgery developed surgical site infection due to delayed ambulation[17]. Moreover, the use of extensive resources and heath costs due to delayed ambulation are high[33].

Patient characteristics, such as age, comorbidities, educational status, residence, and occupation, may influence the timing of early ambulation[34]. Surgical factors, including the type of abdominal surgery, reason of surgery (diagnosis), surgical approach (open or minimally invasive), and intraoperative factors, such as vital signs and duration of surgery, may also impact the initiation of early ambulation[22, 35]. Furthermore, perioperative interventions, such as pain management strategies and the implementation of ERAS protocols, may play a role in facilitating or delaying early ambulation.

Despite the recognized advantages of early ambulation, there exists considerable variability in the time taken by patients to initiate ambulatory activities and low adherence to it after major abdominal surgery. This variability prompts a critical examination of the factors influencing the timing of early ambulation and their impact on overall recovery. More interestingly, there is no any study that assessed the time to early ambulation and its predictors among patients undergoing abdominal surgery in Ethiopia. Thus, this study was aimed to assess the time to early ambulation and its predictors among patients undergoing abdominal surgery in East Gojjam Zone Public Hospitals, Northwest Ethiopia.

## Methods and Materials

### Study area and Period

The study was conducted in East Gojjam Zone Public Hospitals. East Gojjam Zone has eleven public hospitals, including one comprehensive specialized hospital, one general hospital, and nine primary hospitals. These are namely Debre Markos Comprehensive Specialized Hospital, Dejen Primary Hospital, Bichena Primary Hospital, Motta General Hospital, Yejubie Primary Hospital, Debre Work Primary Hospital, Debre Elias Primary Hospital, Lumamie Primary Hospital, Bibugn Primary Hospital, Shebel Berenta Primary Hospital, and Merto-Lemariam Primary Hospital. The study was conducted from June 1^st^ 2024 to December 1^st^ 2024.

### Study design

The study employed a prospective follow up study.

### Source population

All admitted patients undergoing abdominal surgery in East Gojjam Public Hospitals were source population.

### Study population

All admitted patients undergoing abdominal surgery in East Gojjam Public Hospitals during the data collection period were study population.

### Eligibility criteria

The study included admitted adult patients (aged 18 years or older) undergoing major abdominal surgery. On the other hand, patients with significant mobility limitations, and transfer from other health institutions after abdominal operation done were excluded from the study. Additionally, clients undergoing caesarian section for delivery were excluded from the study.

### Sample size determination

The sample size was calculated using Epi Info version 7 by considering proportion of early ambulation in the previous studies in both exposed and unexposed group, power 80%, 5% margin of error, and 95% confidence level (Table 1).

**Table 1:** sample size for a study on time to early ambulation and its predictors among patients undergoing abdominal surgery in East Gojjam Zone Public Hospitals, 2024.

| Factor | Odds ratio | % Outcome in unexposed | % Outcome in exposed | Power | Sample size | Reference |
| --- | --- | --- | --- | --- | --- | --- |
| Being male | 0.56 | 58% | 43.6 | 80% | 404 | [28] |

By adding 10% non-response rate, the final sample size was 444.

### Sampling procedure

Systematic sampling technique was used in all hospitals to select study participants. Initially, the total sample size was proportionally allocated to each hospital by using the previous number of patients underwent abdominal surgery at each hospital. Then, the k value was calculated by dividing the number of study population by sample size which was 2 to select the study participants in every k value. From Yejubie Primary Hospital and Debre-Elias Primary Hospital, 26 participants were selected from each. From Dejen Primary Hospital, 30 participants were selected, and from Motta General Hospital, 47 participants were included. From Bibugn Primary Hospital and Shebel Primary Hospital, 23 participants were selected from each. Additionally, 156 participants were selected from Debre Markos Comprehensive Specialized Hospital, 24 from Debrework Primary Hospital, 36 from Bichena Primary Hospital, 24 from Mertole Le-Mariam Primary Hospital, and 29 from Lumamie Primary Hospital (Figure 1).

**Figure 1:**
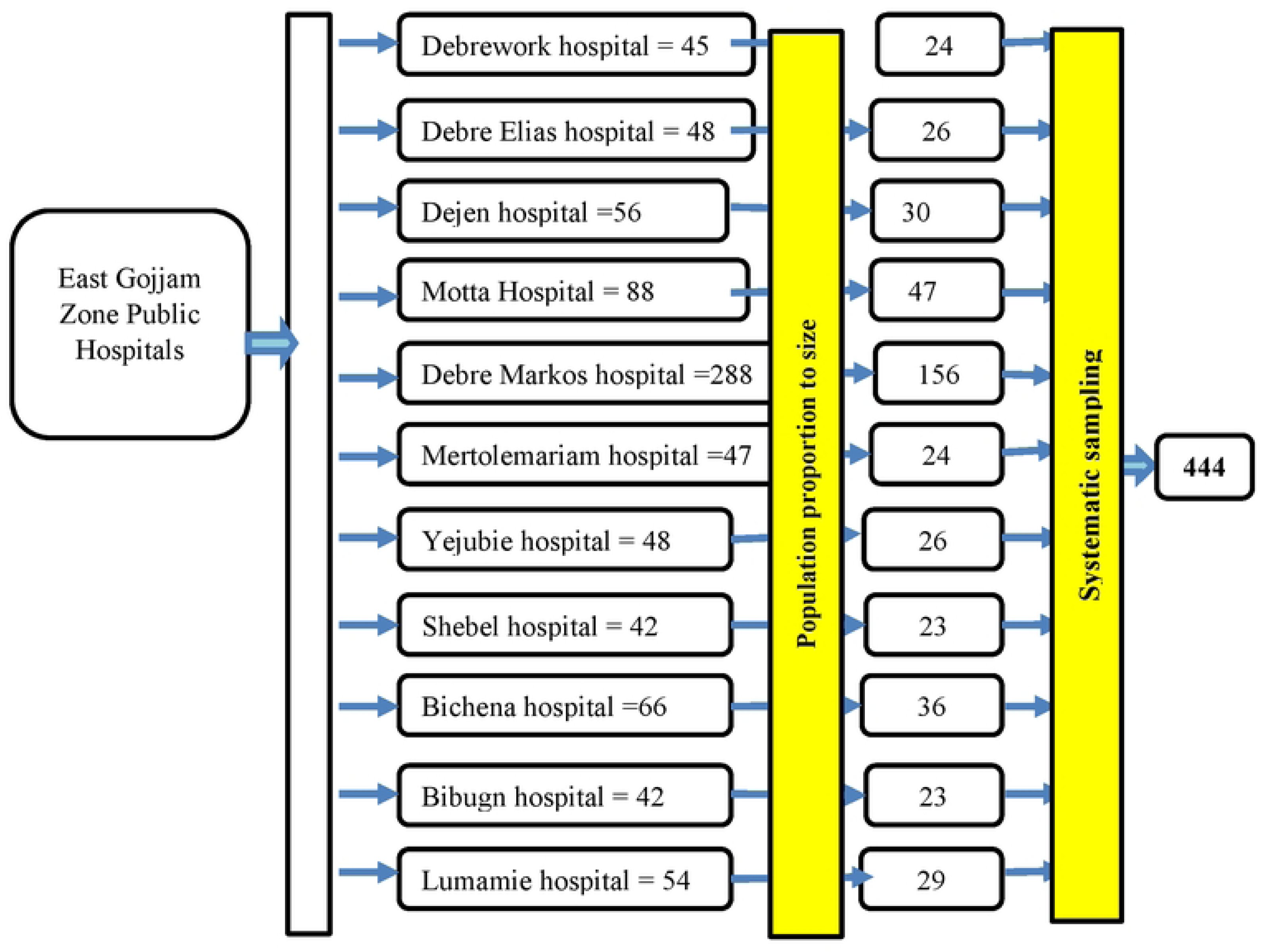
Sampling procedure for time to early ambulation and its predictors among admitted patients undergoing abdominal surgery in East Gojjam Zone Public Hospitals, Northwest Ethiopia, 2024/2025.

### Data Collection tool and procedure

Data were collected by using structured questionnaire prepared from different literature[16, 28, 36, 37]. The tool has different components. The first component was socio-demographic characteristics while the second was clinical and surgery related factors. The data were collected by using interview, direct observation of patients, and chart reviewing. Before the data collection, 11 data collectors and 6 supervisors were recruited. The sociodemographic characteristics (sex, age, educational status, marital status, occupation, family support, residence) and some clinical factors (postoperative pain scores scale, complain after surgery) were collected by asking the patients. Clinical and surgery related data (presence of comorbidities, type of surgery, duration of surgery, and type of antipain given were collected by the data collectors from patient’s chart after the surgery was done. The presence of colostomy, and the presence of catheter were observed directly. Patients were followed for 24 hours, and the time of initiation of walking and movement, and the starting time of feeding and its type were recorded by the patients’ attendants in the absence of the data collectors. Additionally, postoperative vital signs were followed for 24 hours and the average score was recorded. All part of the questionnaire were prepared in English version initially and translated into Amharic then back to English to check their consistency.

### Operational definition

**Time to Early Ambulation**: the duration (in hours) from the end of surgery to the initiation of walking and movement by the patient.

**Early ambulation**: initiating walking and movement by the patient in the day of surgery (within 24 hours)[11, 38].

**Event**: initiation of walking and movement by the patient himself/herself after abdominal surgery within 24 hours.

**Censored**: patient who cannot initiate walking or movement within 24 hours of the surgery, or patients who are moving before completion of the study from the hospitals to other health institutions for care and treatment.

**Follow-up**: Patients will be followed until **24** hours after abdominal surgery.

### Variables

#### Dependent variable

- Time to early ambulation (hours)

#### Independent variables

- Sociodemographic characteristics: sex, age, educational status, residence, occupation, marital status, support from family
- Clinical and surgery related characteristics: presence of comorbidities, reason of surgery (diagnosis), type of surgery, duration of surgery, postoperative pain scores, type of antipain, intraoperative vital signs (systolic blood pressure, diastolic blood pressure, pulse rate, respiratory rate and temperature), postoperative vital signs (systolic blood pressure, diastolic blood pressure, pulse rate, respiratory rate and temperature), recent hemoglobin, complain after surgery, presence of colostomy, presence of catheter, perioperative blood loss, postoperative nutritional status (time to start feeding, type of feeding)

### Data quality control

Prior to data collection, 2 days training was given to data collectors and supervisors on questionnaire content, methods of data collection, and ethical concerns. To improve the quality of data, pretest was done on five percent (5%) of questionnaires before two weeks of actual data collection at Dembecha hospital. After reviewing the result of the pretest, rearrangement of the questionnaire was performed for clarity and completeness of the questionnaire. Continuous supervision of data gathering and daily checking of the collected data were performed by supervisors and principal investigator. After data collection, data verification was conducted visually and possible corrections were made by cross-checking the questionnaire.

### Data processing and analysis

The data were initially coded, checked in Kobo collect and then exported to STATA version 17 for cleaning and statistical analysis. The survival function was generated by using the Kaplan Meier curve. Log-Rank test was performed to see the association of each stratum of categorical variables with the survival probability of participants. The median time to early ambulation was calculated. Before running the cox regression, the Schoenfeld residuals proportional hazard assumption test for each variable was done. P value of the global Schoenfeld residuals proportional hazard assumption test was 0.6223. Moreover, the assumptions of proportion hazard over time were graphically checked. Multicollinearity was checked by using Variance Inflation Factor (VIF) for continuous variables and after creating dummy variables for categorical variables. The highest result of VIF was 3.81. The linear relation of continuous variables was checked by using Martingale residuals. After checking the proportional hazard assumption, multicollinearity, linearity of each continuous variables with the outcome variable, cox proportional regression analysis model was enrolled. The fitness of the model was checked by using Cox – Snell test. The. Initially, bivariable cox regression analysis was done to select variables for multivariable analysis. Variables with a P value less than 0.2 in the bivariable analysis were selected as candidates for the multivariable cox regression analysis model. The level of statistical significance will be declared at a P-value less than 0.05. An adjusted hazard ratio with its 95% CI will be used to show the strength and direction of association between each explanatory variable and the outcome variable.

## Result

### Sociodemographic characteristics of patients undergoing abdominal surgery

A total of 444 patients undergoing abdominal surgery were followed. Among these, 131 (29.5%) were female, and 272 (61.3%) were married. The median age of the study participants was 35 years (IQR: 21). Regarding educational status, 161 (36.3%) were unable to read and write. Nearly half of the participants, 221 (49.8%), were farmers, and 239 (53.8%) lived in rural areas. Additionally, 51 (11.5%) did not receive support from their family or attendants during their admission (Table 2).

**Table 2:**
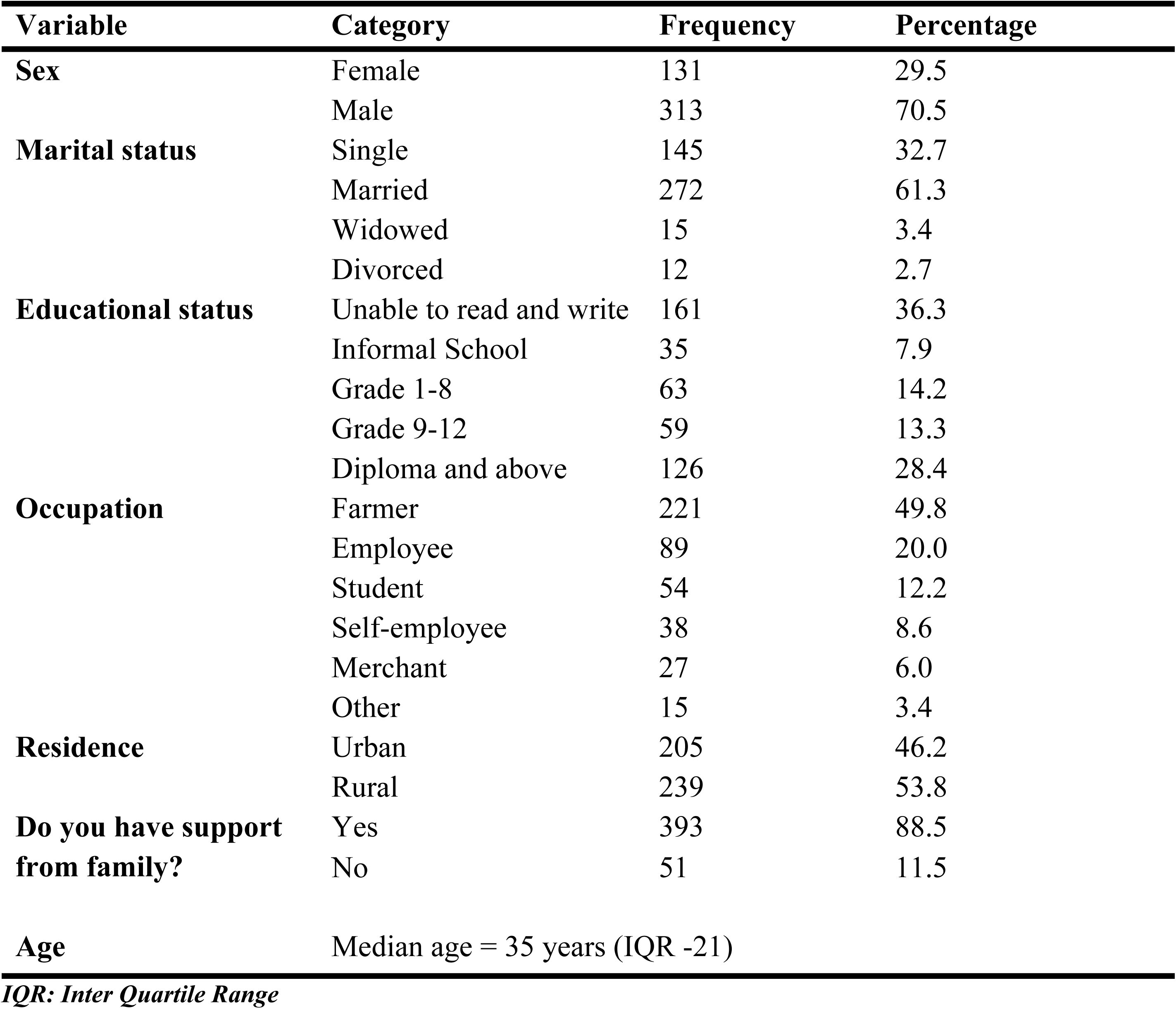
sociodemographic characteristics of patients undergoing abdominal surgery in East Gojjam Zone Public Hospitals, Northwest Ethiopia, 2025 (N-444)

| Variable | Category | Frequency | Percentage |
| --- | --- | --- | --- |
| **Sex** | Female | 131 | 29.5 |
|  | Male | 313 | 70.5 |
| **Marital status** | Single | 145 | 32.7 |
|  | Married | 272 | 61.3 |
|  | Widowed | 15 | 3.4 |
|  | Divorced | 12 | 2.7 |
| **Educational status** | Unable to read and write | 161 | 36.3 |
|  | Informal School | 35 | 7.9 |
|  | Grade 1-8 | 63 | 14.2 |
|  | Grade 9-12 | 59 | 13.3 |
|  | Diploma and above | 126 | 28.4 |
| **Occupation** | Farmer | 221 | 49.8 |
|  | Employee | 89 | 20.0 |
|  | Student | 54 | 12.2 |
|  | Self-employee | 38 | 8.6 |
|  | Merchant | 27 | 6.0 |
|  | Other | 15 | 3.4 |
| **Residence** | Urban | 205 | 46.2 |
|  | Rural | 239 | 53.8 |
| **Do you have support from family?** | Yes | 393 | 88.5 |
|  | No | 51 | 11.5 |
| **Age** | Median age = 35 years (IQR -21) |  |  |

*IQR: Inter Quartile Range*

### Clinical and surgery related characteristics of patients undergoing abdominal surgery

From the total of 444 patients undergoing abdominal surgery, 9(2%) of them had history of comorbidity. One third, 150(33.8%) of them were undergoing abdominal surgery because of appendix case such as appendicitis, appendiceal abscess, and perforated appendix, and 93(21%) were operated due to intestinal obstruction. Moreover, 62(14%) of them were undergoing abdominal surgery as a result of injury. Regarding to complain occurred after surgery, 21(4.7%) and 4(0.9%) were unable to respond and desaturated respectively. Forty-eight (10.8%) and 287(64.6%) had colostomy and catheter respectively. Furthermore, 51(11.5%) took tramadol for antipain. Three of the total study participants took water for the first time after surgery. The median time for the duration of surgery was 1 hour (IQR: 1.25) and the median amount of estimated blood loss during surgery was 800ml (IQR: 200). The median time to start feeding after surgery was 13 hours (IQR: 9). The median postoperative diastolic blood pressure was 71mmHg (IQR-13) and the postoperative respiratory rate 20 breath per minute (IQR-0). In addition to this, the postoperative body temperature was 36°C (Table 3).

**Table 3:**
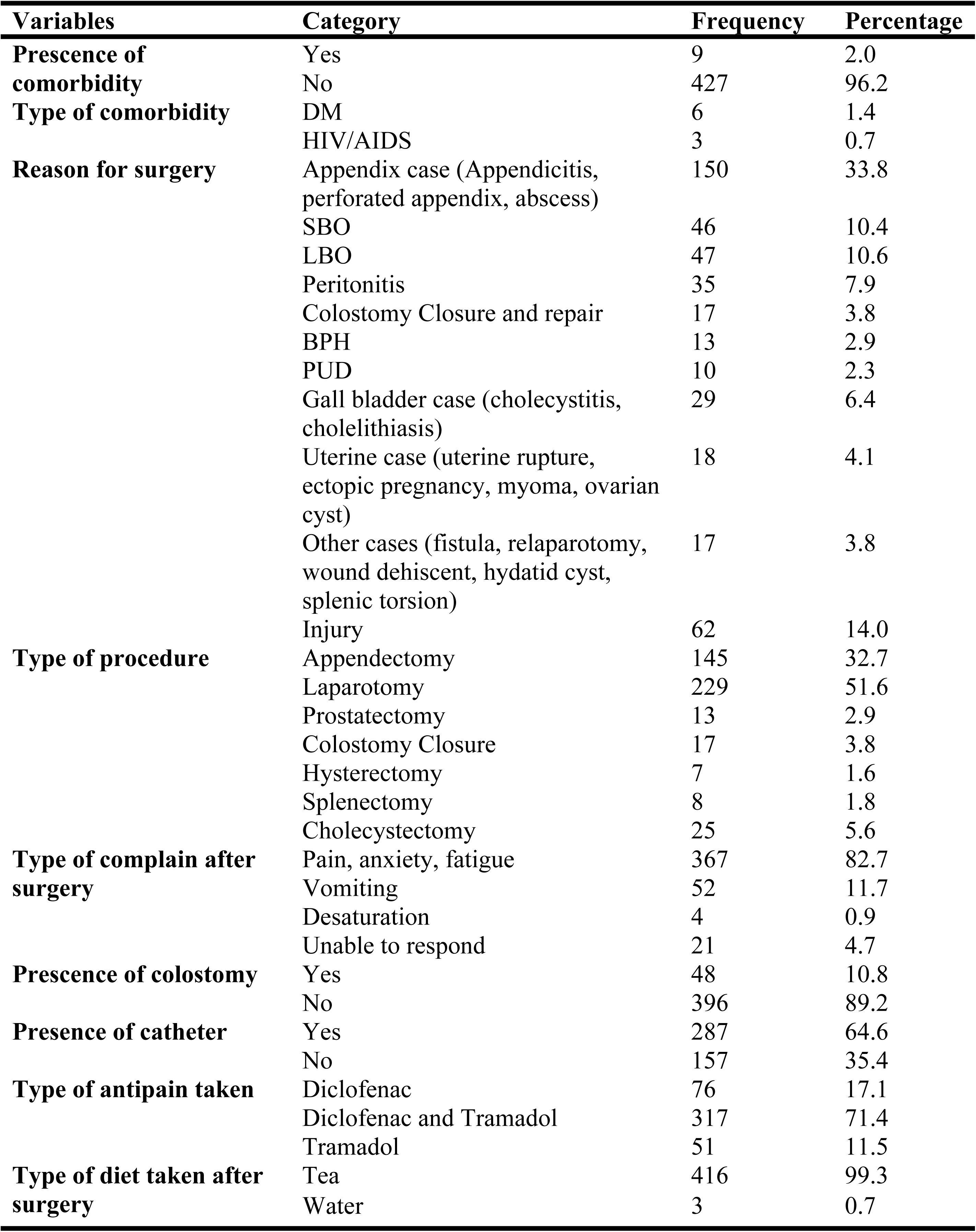
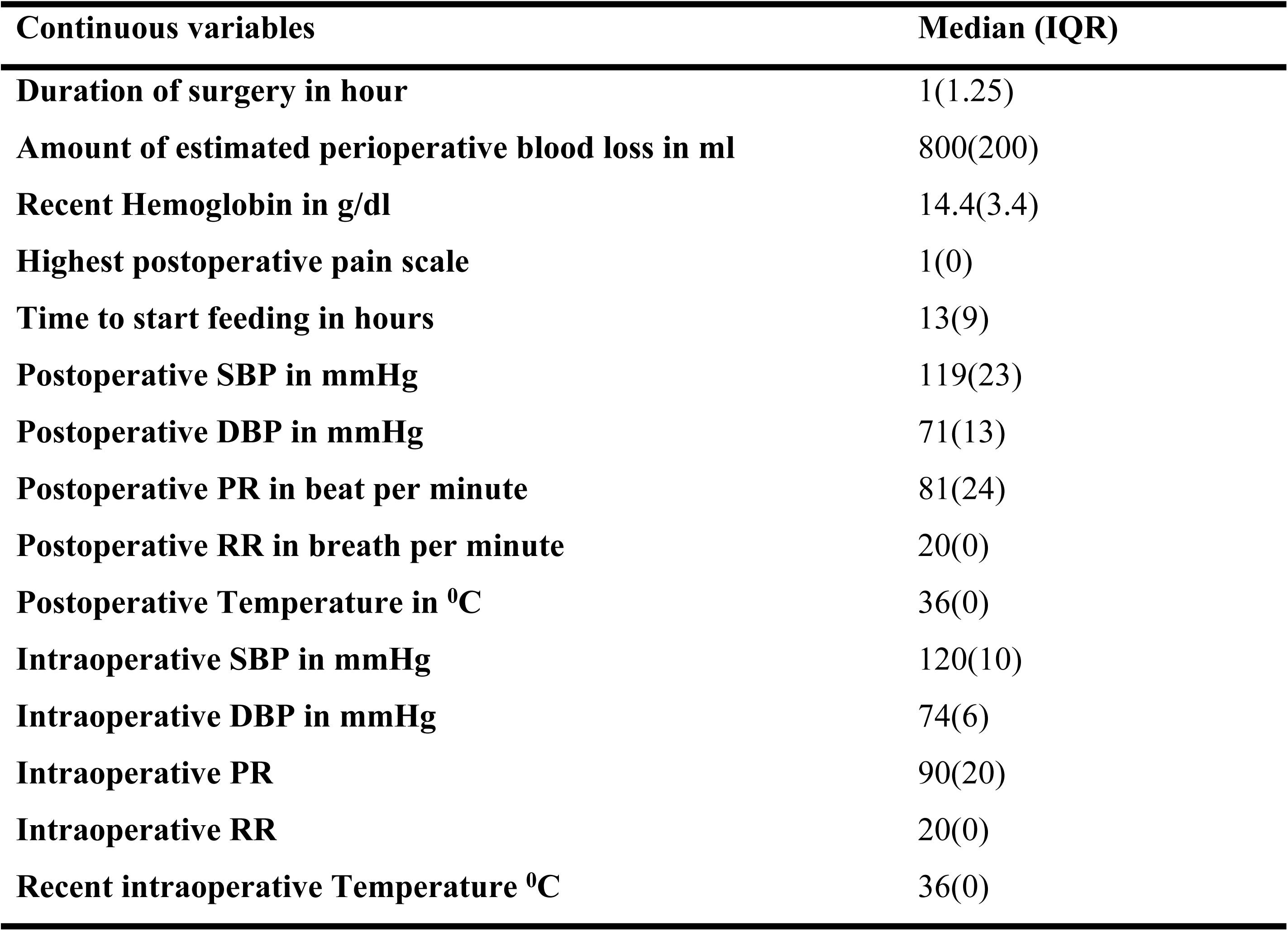
Clinical and surgery related characteristics of patients undergoing abdominal surgery, 2025.

### Time to Early Ambulation

A total of 444 patients undergoing abdominal surgery were followed for 6354.8 hours, with the minimum follow up time was 3 hours and the maximum was 24 hours. Among these, 358 (80.6%) were ambulated within 24 hours. The incidence rate of early ambulation among patients undergoing abdominal surgery was 0.0564 per patient-hour (95% CI: 0.0507 – 0.0626). This indicates that, on average, approximately 5.64 patients achieve early ambulation for every 100 patient-hours of follow-up. The median time to early ambulation among patients undergoing abdominal surgery was 13 hours, meaning that 50% of patients achieved early ambulation within 13 hours (Figure 2).

**Figure 2:**
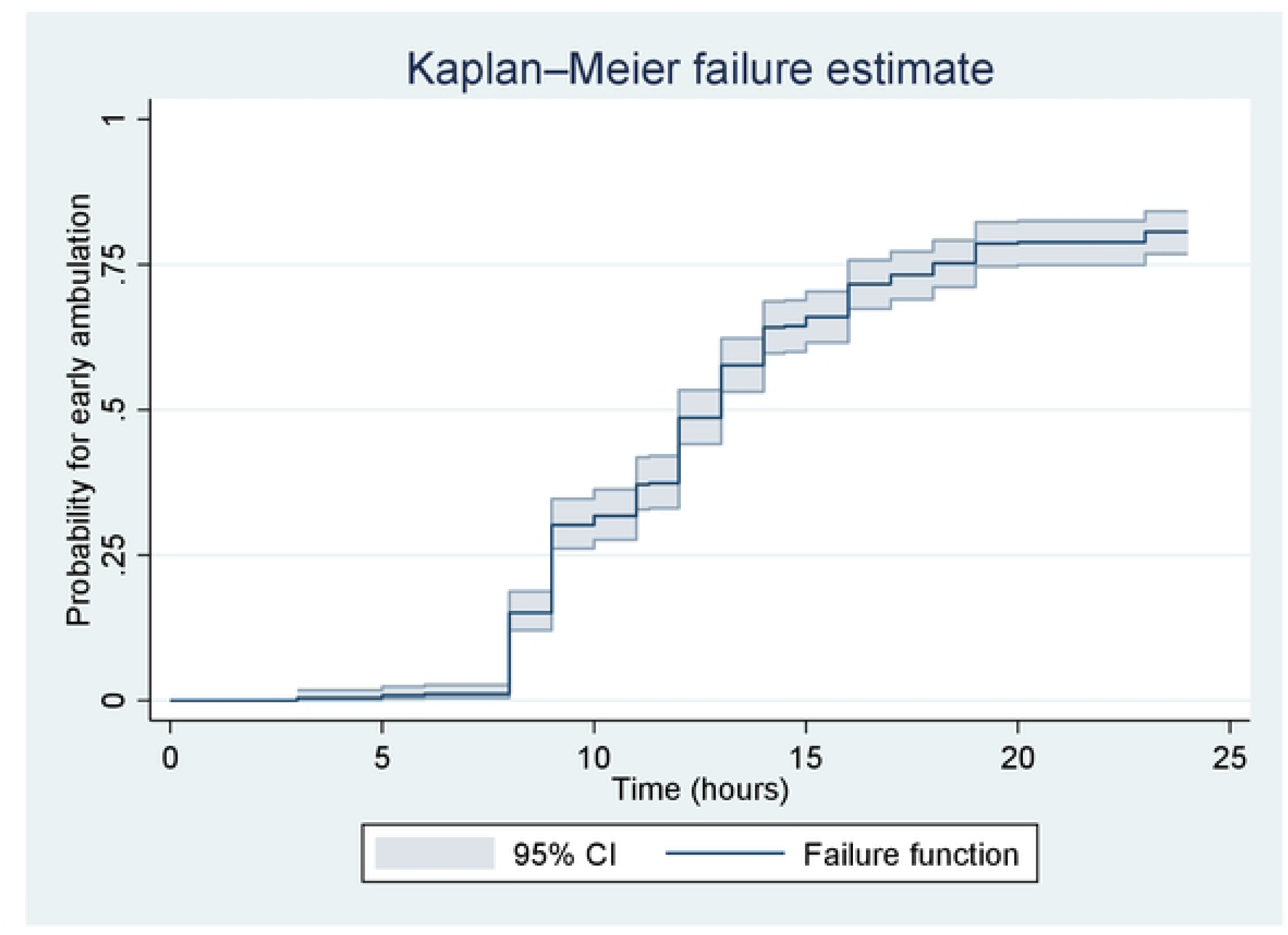
The overall Kaplan-Meier estimate for time to early ambulation, 2025.

### Survival function among categorical variables

The difference in time to early ambulation among categorical variables was assessed graphically using Kaplan-Meier curves and statistically using the Log-rank test. In the Kaplan-Meier graph, the lower curve indicates those group of people who took a longer time to develop the event. The time to early ambulation was found to be statistically different between male and female patients (p < 0.001) (Figure 3). Additionally, significant differences in time to early ambulation were observed based on educational status (Figure 4), marital status (Figure 5), type of antipain medications (Figure 6), presence of a catheter (Figure 7), and the presence of support level (Figure 8). The median time to early ambulation varied by surgical case type: appendix cases had the earliest median time (9 hours), while colostomy cases (18 hours) and injured cases had delayed ambulation times. In terms of incidence rates, appendix cases and BPH (benign prostatic hyperplasia) cases had higher incidence rates, suggesting that a greater number of these cases were ambulated per 100 patient-hours of follow-up (Table 4).

**Figure 3:**
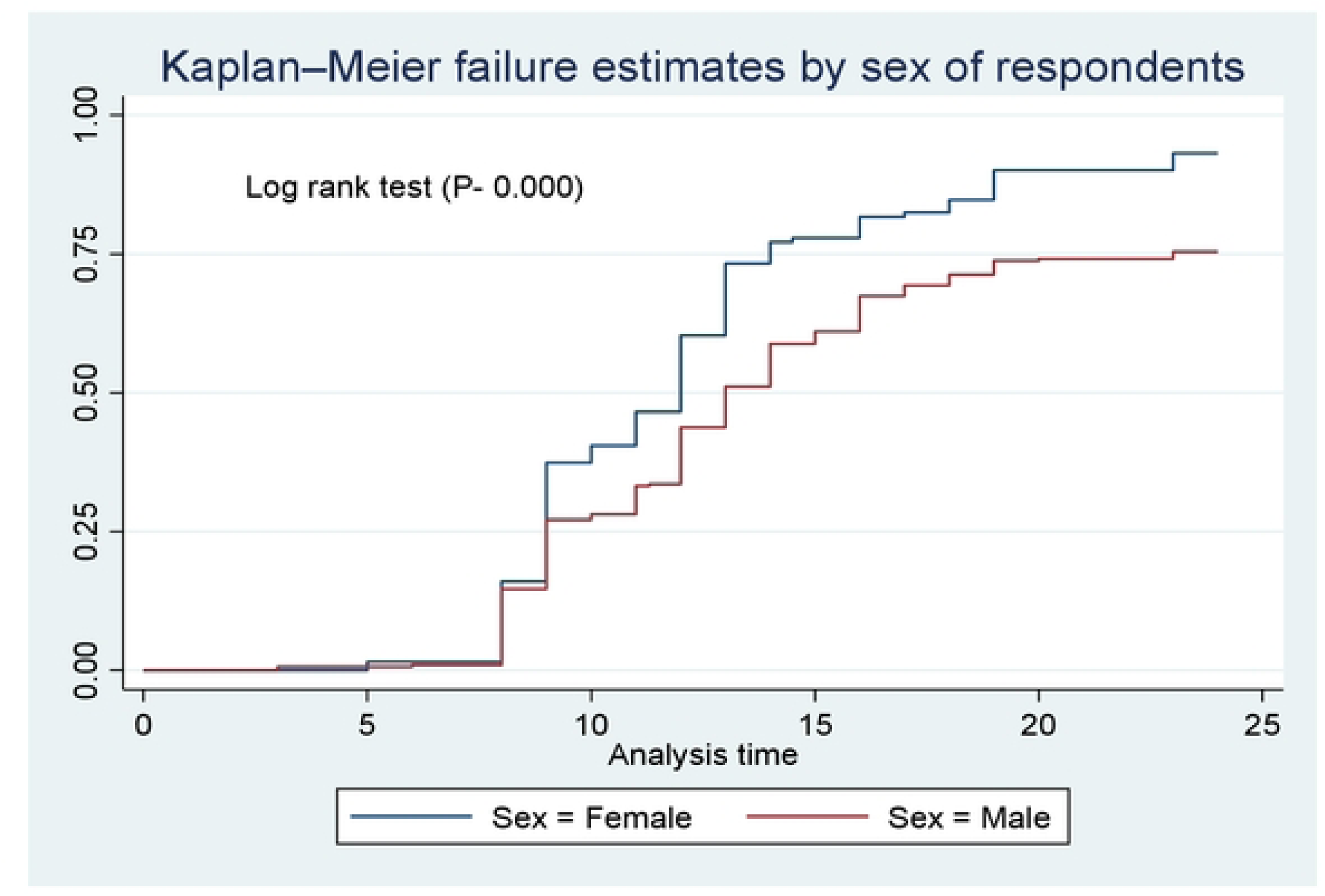
The Kaplan-Meier curve of time to early ambulation among female and male patients, 2025.

**Figure 4:**
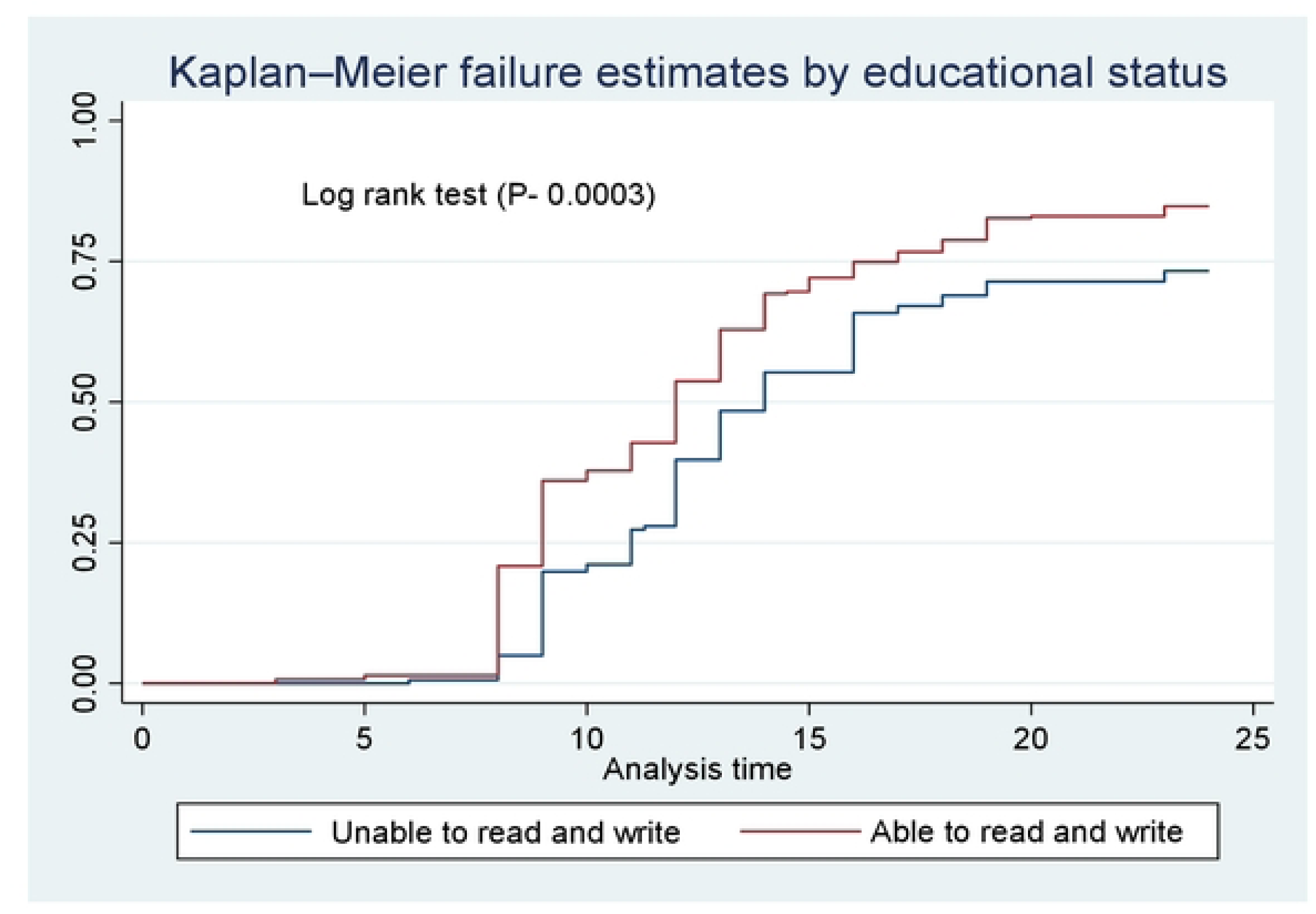
The Kaplan-Meier curve of time to early ambulation by educational status, 2025.

**Figure 5:**
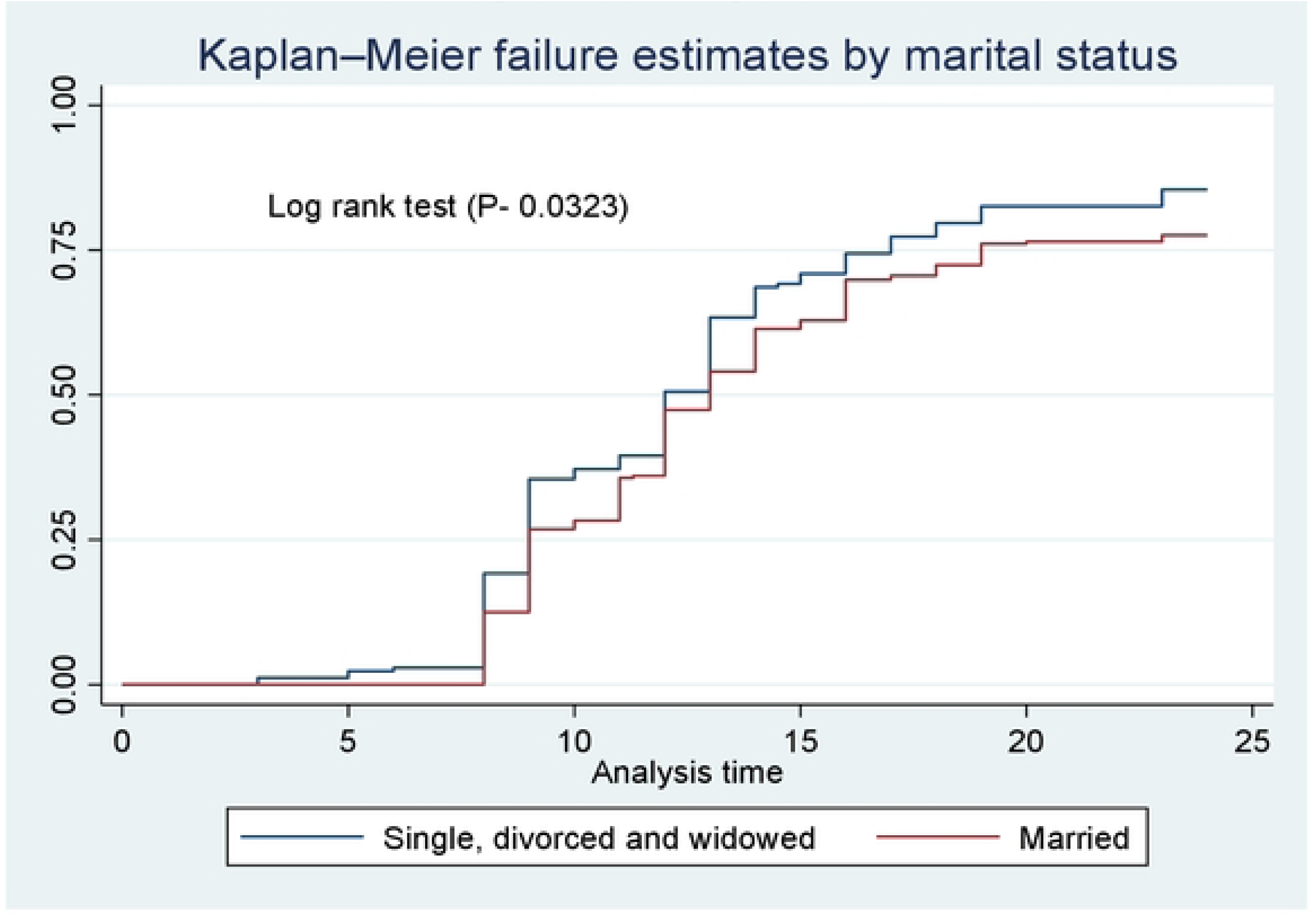
The Kaplan-Meier curve of time to early ambulation by marital status, 2025.

**Figure 6:**
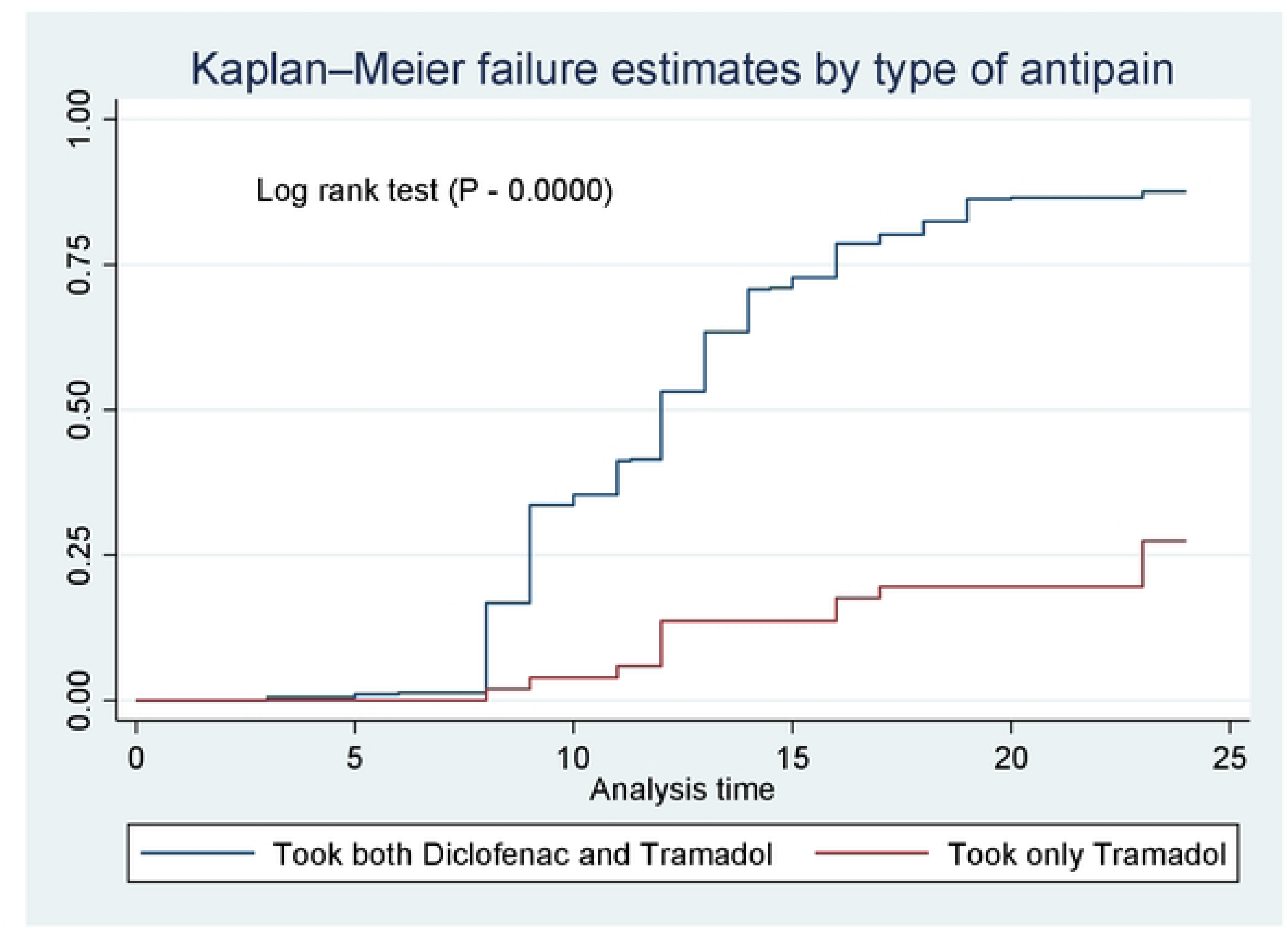
The Kaplan-Meier curve of time to early ambulation by the type of antipain, 202S.

**Figure 7:**
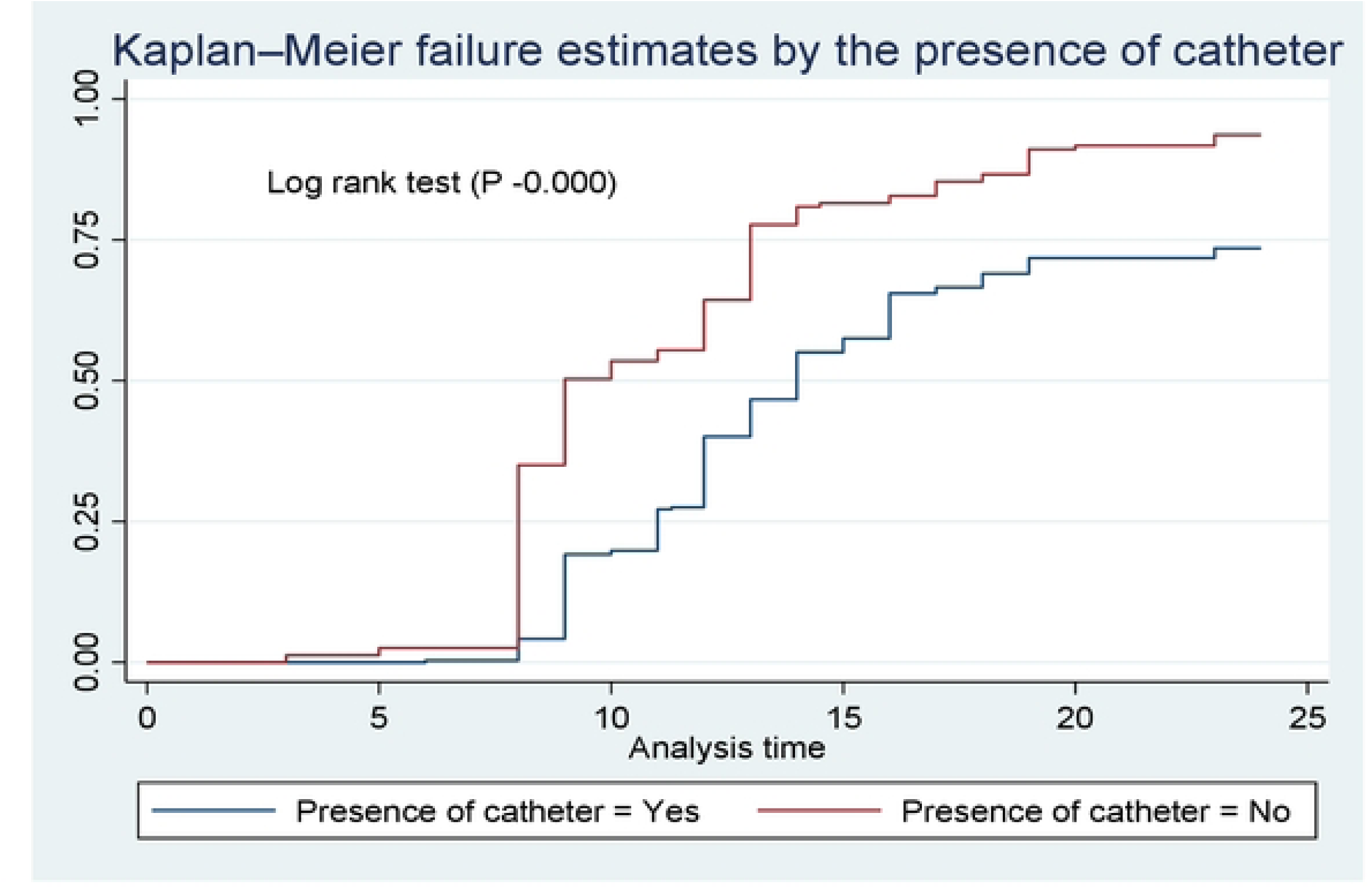
The Kaplan-Meier curve of time to early ambulation by the presence of catheter, 202S.

**Figure 8:**
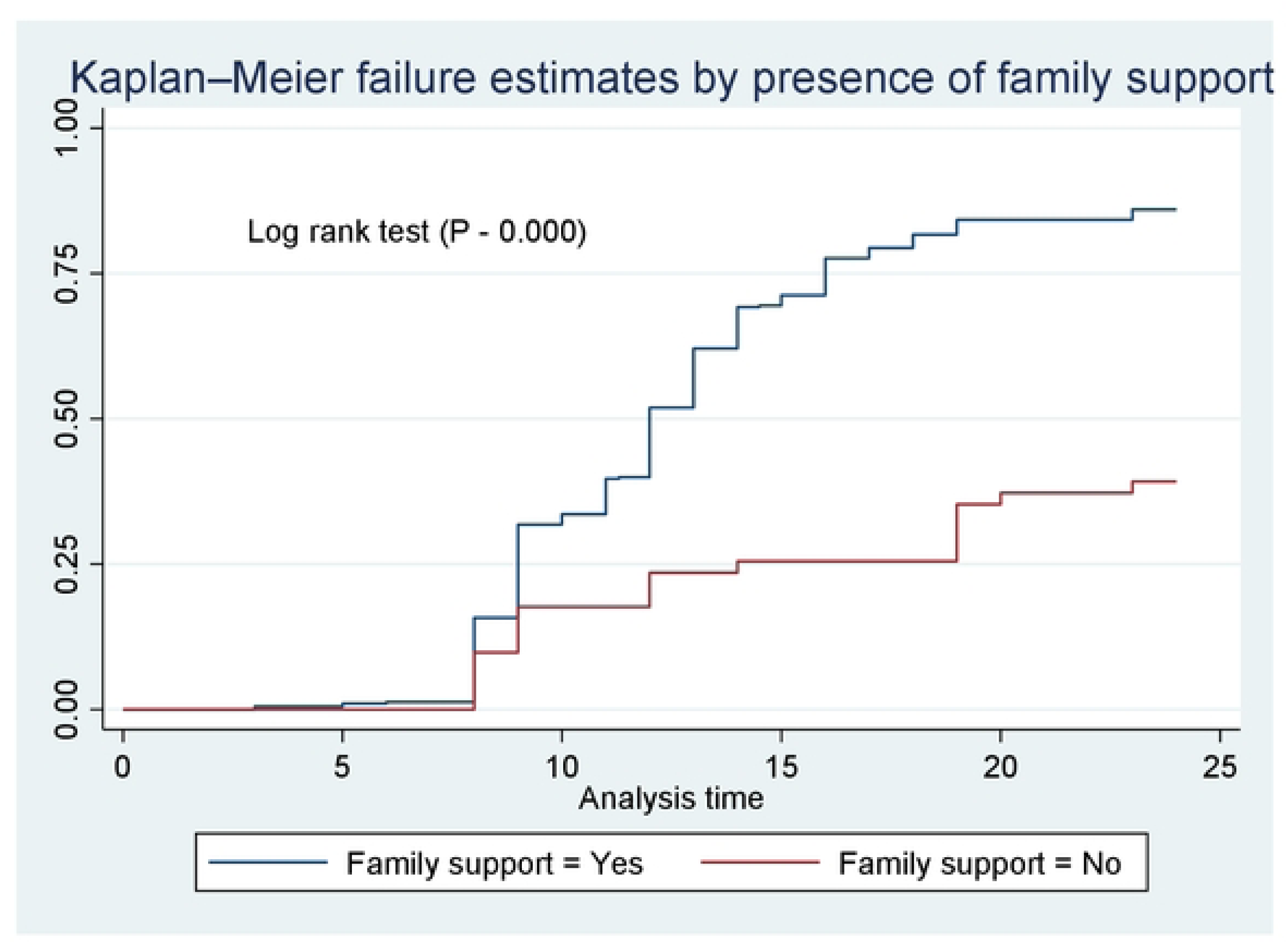
The Kaplan-Meier curve of time to early ambulation by the presence of family support, 2025.

**Table 4:** incidence and median time of early ambulation by reason of surgery (DX), 2025.

| Reason for surgery (DX) | Incidence rate | Median time for early ambulation |
| --- | --- | --- |
| Appendix case (Appendicitis, perforated appendix, appendiceal abscess) | 0.091 | 9 |
| SBO | 0.050 | 14 |
| LBO | 0.049 | 15 |
| Peritonitis | 0.067 | 13 |
| Colostomy Closure and repair | 0.028 | 18 |
| BPH | 0.084 | 12 |
| PUD | 0.069 | 11 |
| Gall bladder case (cholecystitis, cholelithiasis) | 0.062 | 13 |
| Uterine case (uterine rupture, ectopic pregnancy, myoma, ovarian cyst) | 0.075 | 12 |
| Other cases (Fistula, relaparotomy, wound dehiscent, hydatid cyst, splenic torsion) | 0.046 | 14 |
| Injury | 0.015 | - |

Note: “-“ indicates that only 25% of injured cases were ambulated. The rest 75% injured cases were censored.

### Predictors to Time to Early Ambulation among patients undergoing abdominal surgery

Variables which fulfilled the assumptions of cox proportional hazard model were entered to the bivariable cox proportional hazard regression model. Then variables whose P value was less than 0.2 were entered to multivariable cox proportional hazard regression. These variables which were entered to multivariable were sex, age, marital status, presence of family support, type of anti-pain taken, presence of catheter, estimated blood loss, postoperative diastolic pressure, postoperative respiratory rate, intraoperative respiratory rate, pulse rate and temperature. However, only age, presence of catheter, type of antipain taken, postoperative diastolic blood pressure, postoperative respiratory rate, intraoperative pulse rate and respiratory rate. The model was fitted to variables as the cox Snell residual and the hazard follows the 45^0^ (Figure 9). According to the result, when the age of patients was increased by one year, the risk of early ambulation was decreased by 2% (AHR = 0.98; 95%CI: (0.97-0.99)). The risk to early ambulate among patients who had no catheter in their body tract was 77% higher than patients who had not catheter (AHR= 1.77; 95% CI: (1.37-2.29)). On the other hand, the risk to early ambulate for patients who were taking only tramadol for antipain was 74% less than those who were taking diclofenac (AHR= 0.26; 95% CI: (0.15-0.45)). When the postoperative diastolic blood pressure was increased by 1mmHg, the risk of early ambulation was increased by 1% (AHR-1.01; 95% CI: (1.00-1.02)). When the postoperative respiratory rate was increased by 1breath per minute, the hazard of early ambulation was decreased by 19% (AHR-0.81; 95% CI: (0.75-0.87)). Furthermore, if the intraoperative pulse rate was increased 1 beat per minute, the risk of early ambulation was decreased by 2% (AHR-0.98, 95% CI: (0.97-0.99)). Most interestingly, when the recent intraoperative temperature was increased by 1°C, the risk of early ambulation was increased by 50% (AHR-1.50, 95% CI: (1.11-2.03)) (Table 5).

**Figure 9:**
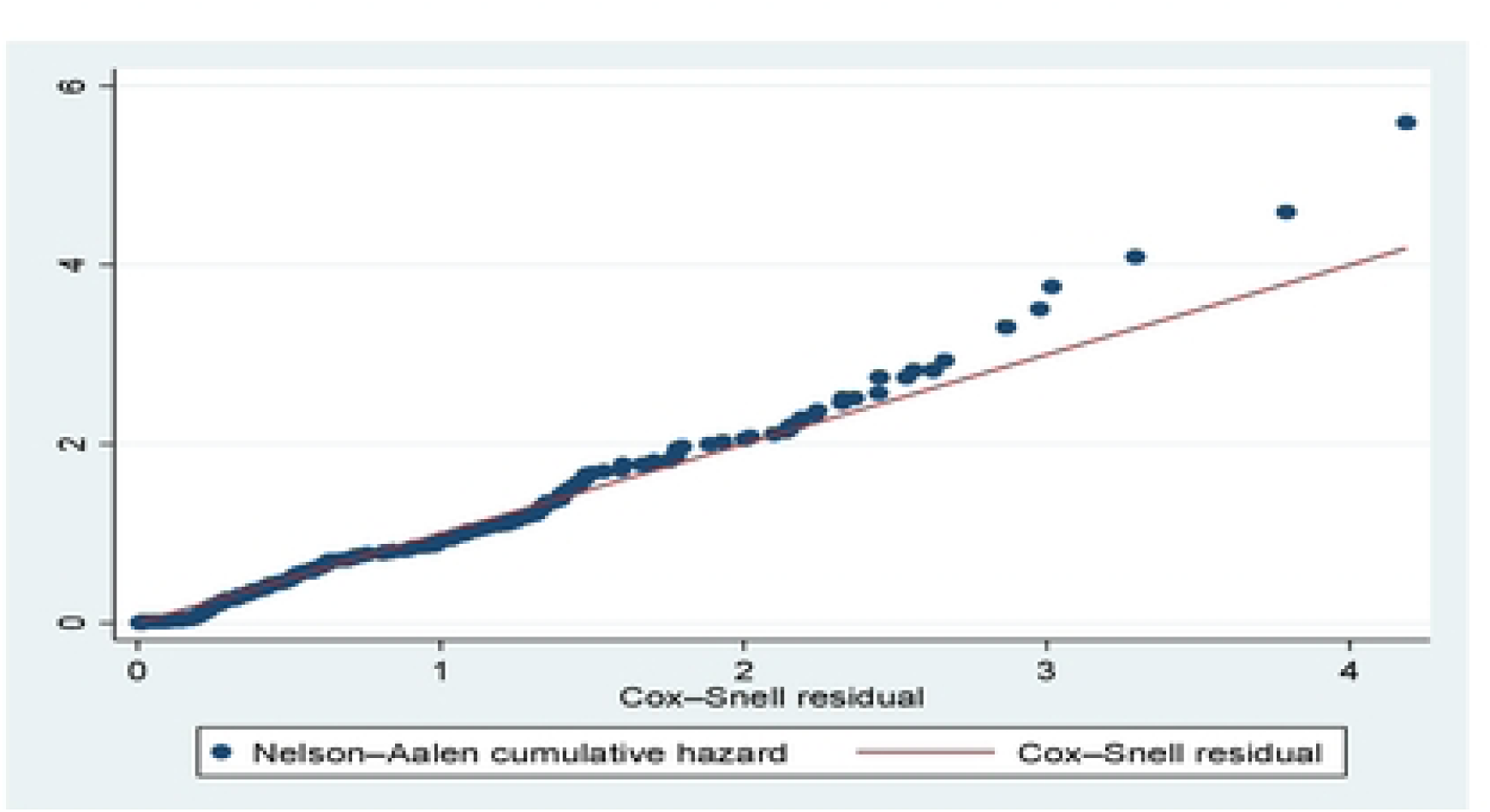
The model fitness test-based cox Snell residual.

**Table 5:** Bivariable and multivariable Cox regression output on the association between time to early ambulation and predictors, 2025 (N= 444)

| Variable | Category | Early ambulation |  | CHR (95% CI) | AHR (95% CI) | P value |
| --- | --- | --- | --- | --- | --- | --- |
|  |  | Ambulated | Censored |  |  |  |
| Sex | Female | 122 | 9 | 1 | 1 | 0.592 |
|  | Male | 236 | 77 | 0.63(0.50-0.78) | 0.94(0.74-1.18) |  |
| Marital status | Single, widowed and divorced | 147 | 25 | 1 | 1 | 0.293 |
|  | Married | 211 | 61 | 0.81(0.65-0.99) | 1.10(0.86-1.42) |  |
| Presence of family support | Yes | 338 | 55 | 3.66(2.32-5.76) | 1.43(0.84-2.43) | 0.192 |
|  | No | 20 | 31 | 1 | 1 |  |
| Presence of catheter | Yes | 211 | 76 | 1 | 1 | **0.000** |
|  | No | 147 | 10 | 2.08(1.68-2.57) | 1.77(1.37- 2.29) |  |
| Type of antipain | Diclofenac | 344 | 49 | 1 | 1 | **0.000** |
|  | Only tramadol | 14 | 37 | 0.16(0.09-0.27) | 0.26(0.15-0.45) |  |
| Age in years |  |  |  | 0.98(0.980-0.99) | 0.98(0.97-0.99) | **0.000** |
| Amount of blood loss in ml |  |  |  | 0.99(0.99-1.00) | 1.00(0.99-1.00) | 0.817 |
| Postoperative DBP in mmHg |  |  |  | 1.01(1.01-1.02) | 1.01(1.00-1.02) | **0.002** |
| Postoperative RR in breath per minute |  |  |  | 0.79(0.76-0.84) | 0.81(0.75-0.87) | **0.000** |
| Intraoperative PR beat per minute |  |  |  | 0.98(0.98-0.99) | 0.98(0.97-0.99) | **0.001** |
| Intraoperative RR in breath per minute |  |  |  | 0.86(0.81-0.90) | 0.99(0.91-1.09) | 0.996 |
| Recent Intraoperative temperature |  |  |  | 2.11(1.66-2.68) | 1.50(1.11-2.03) | **0.004** |

Notes: Schoenfeld residuals proportional hazard assumption test $P = 0.6223$ ; DBP- Diastolic Blood Pressure; PR- Pulse Rate; RR – Respiratory Rate;

## Discussion

Early ambulation among patients undergoing abdominal surgery has a positive impact the prognosis and recovery of the disease condition in addition to preventing the complications. Despite this fact, the median time to early ambulation for patients undergoing abdominal surgery is not known. Hence, this follow up study was determined the median time and the predictors of early ambulation among patients undergoing abdominal surgery at East Gojjam Zone Public Hospitals.

In this study, the cumulative incidence of early ambulation was 80.6% (95% CI: 0.77-0.84). This finding is higher that the findings of a retrospective study conducted in India, 68.2%[22], a prospective cohort study which was conducted in Australia and New Zealand among patients admitted in intensive care unit, 35.5% [39], a prospective study which was conducted in factors impacting early mobilization according to the Enhanced Recovery After Surgery guideline following gastrointestinal surgery in China, 53.6% [37]. On the contrary, the finding of this study is lower than a quasi non randomized trial conducted in Turkey, 100% of the study participants initiated early mobilization within 24 hours of the surgery[25]. The possible justification for this discrepancy could be the difference in study population and the study design. Unlike to this study which included different cases, some of the studies were done among specific cases such as colorectal cancer cases and critically ill patients [39]. The study designs used for the previous studies were retrospective [22], and quais experimental study [25]. Additionally, in this study, the incidence rate for early ambulation was 5.64% (95% CI: 0.0507 – 0.0626) per 100 patient-hours follow up. The median time for early ambulation for patients undergoing abdominal surgery was 13 hours although it differs from diagnosis to diagnosis. For example, patients with appendix case (appendicitis, appendiceal abscess, perforated appendix) had ambulated early compared to others. This finding is in line as to the guideline of Enhanced Recovery After Surgery.

The risk of early ambulation is decreased when the age of patients is increased. This finding is supported by a study done on association between delayed ambulation and increased risk of adverse events after lumbar fusion surgery in elderly patients[40]. The possible justification for this might be reduced muscle strength and endurance, greater pain sensitivity and decreased cardiopulmonary reserve while age is increased[41]. In addition to this, aged patients will have been dependent on care givers, and may have cognitive issues that hinder compliance of postoperative instructions.

The association between antipain and early ambulation was also observed. Patients who took only tramadol for pain management have less risk to early ambulation. This occurs due to that tramadol causes drowsiness, dizziness and confusion[42]. Moreover, tramadol results in nausea and vomiting[43] which delayed initiation of early ambulation.

The other predictor that affects the incidence of early ambulation was the presence of catheter. In this study, the risk to ambulate early among patients who had not catheter was higher than patients who had it. This is supported by a prospective study done on factors impacting early mobilization according to the Enhanced Recovery After Surgery guideline following gastrointestinal surgery in China[34]. The possible justification for this might be the discomfort of catheter during ambulation. When the catheter is inserted somewhere in the body tract, it would irritate and embarrassed the ambulating patient. Furthermore, patients may afraid the dragging and fall of catheter. As a result, patients may avoid walking and early ambulation.

More interestingly, this study found that increment of postoperative diastolic blood pressure increases the risk of early ambulation. This implies that when the postoperative DBP is increased, ambulation will be early. This occurs due to that higher diastolic blood pressure results in better perfusion of oxygen and nutrient to muscles and tissue. Additionally, since moderate increment of blood pressure decreases the sensitivity of pain receptors[44], the risk of early ambulation will be increased. Nevertheless, there are other evidences that show higher diastolic blood pressure increases the occurrence of dizziness and workload of heart [45, 46] which in turn results in fatiguability and delayed ambulation.

This study reveals that the hazard of early ambulation is decreased while the postoperative respiratory rate is increased. Increment of respiratory rate after surgery is a sign of pain, hypoxia, anxiety, postoperative complications such as atelectasis and metabolic acidosis[47]. When these conditions occurred, patients feel fatigue and their desire to ambulate early is decreased.

Moreover, the hazard of early ambulation is decrease when the intraoperative pulse rate is increased. The possible justification for this might be hemodynamic instability. A high pulse rate may indicate hypovolemia, hypotension, or poor cardiac output[48], leading to dizziness, weakness, and an increased risk of falls during ambulation.

The other predictor that enhances early ambulation is elevation of temperature either at preoperative or intraoperative phase. In this study, the risk of early ambulation was increased when the intraoperative temperature is increased. This may be due to vasodilation effect of hyperthermia. A higher temperature leads to vasodilation, enhancing blood flow to muscles and tissues[49], which may reduce stiffness and promote early mobility. Additionally, a warmer body temperature supports enzymatic and cellular functions, aiding in faster wound healing and muscle recovery, which encourages early mobilization. Warmed muscles are more flexible and less prone to stiffness[50], making it easier for patients to move soon after surgery.

### Limitation of study

Although this study has its own strengths, it has also limitations. The study was done among all types of patients undergoing abdominal surgery. Thus, it is difficult to generalize for a specific disease condition. Additionally, the sample size could be enough since the sample size was not calculated based on the previous follow-up study.

## Conclusion

The median time to early ambulation among patients undergoing abdominal surgery was 13 hours although it differs from case tot case. A significant number patients undergoing abdominal surgery were ambulated as to the Enhanced Recovery After Surgery guideline. Increased age, not having catheter, taking tramadol for antipain, postoperative diastolic blood pressure and respiratory rate, intraoperative pulse rate and temperature were predictors of time to early ambulation. Health care providers, particularly nurses and surgeons, should actively promote and facilitate early ambulation within 13 hours following abdominal surgery. Furthermore, the use of alternative analgesics to tramadol is recommended for postoperative pain management. Special consideration should be given to patients who are aged and those with urinary catheters, as they may require additional support during the initiation of ambulation. Nurses and surgeons must also monitor postoperative diastolic blood pressure, as well as respiratory complications such as oxygen desaturation and fatigue. Intraoperative hemodynamic stability, including pulse rate and body temperature, should be maintained through coordinated efforts between anesthesia professionals and the other surgical team. It is better to have a guideline at hospitals that enhance early ambulation among patients undergoing surgery at their median time of early ambulation. Researchers better to investigate the predictors early ambulation by qualitative study. Moreover, it is better to study the median time of early ambulation for each case by including variables such as body mass index and type of anesthesia drugs.

## Data Availability

All relevant data are within the manuscript and its supporting information files.

## Abbreviation and Acronyms

AHR: Adjusted Hazard Ratio
DBP: Diastolic Blood Pressure
ERAS: Enhanced Recovery After Surgery
VIF: Variance Inflation Factor
SBP: Systolic Blood Pressure
PR: Pulse Rate
RR: Respiratory Rate

## Declarations

### Ethical consideration

Data collection was started after the study was approved by the Debre Markos University College of Medicine and Health Sciences Institutional Research Ethical Review Committee (IRERC) with the reference number: RCSTTD/353/01/16 on the date of 30/05/2024. After that the permission letter was obtained from hospitals. Written informed consent was taken from all study participants, with full right to refuse participating in the study. The confidentiality of the records was preserved throughout the study. Respondents’ responses excluded the names and identifiers of study.

### Consent for publication

“Not applicable”

### Availability of data and materials

All relevant data are within the manuscript and its Supporting Information files.

### Author’s contribution

**AE and TL** conceived and designed the study, performed analysis and interpretation of data. **TW, AK, BA, and SA** were involved in the data collection and supervision of work. **TA** and **AG** were involved in the data analysis and interpretation. **AS** and **BZ** drafted the manuscript. All authors read and approved the final Manuscript.

## Acknowledgement

We, the authors, want to acknowledge our university, Debre Markos University for giving this chance. We also have a gratitude for Research, Community Service and Technology Transfer Directorate of College of Medicine and Health Science for guiding us throughout the process. Next to this, we are deeply indebted to acknowledge the data collectors and study participants for giving their time.

